# EFFICACY OF SIBUTRAMINE AND TOPIRAMATE COMBINATION IN WEIGHT LOSS: RETROSPECTIVE COHORT STUDY ON BRAZILIAN ADULTS

**DOI:** 10.1101/2024.05.25.24307923

**Authors:** Heloisa Harumi Yamamoto, Thiago Fraga Napoli, João Eduardo Nunes Salles

**Affiliations:** ¹ Faculdade de Ciências Médicas da Santa Casa de São Paulo (FCMSCSP), Department of Internal Medicine, Discipline of Endocrinology, São Paulo, SP, Brazil; ² Faculdade de Ciências Médicas da Santa Casa de São Paulo (FCMSCSP), Department of Internal Medicine, Discipline of Endocrinology, State Government Employee Medical Assistance Institute (IAMSPE), São Paulo, SP, Brazil; ³ Vassouras University, Department of Internal Medicine, Discipline of Endocrinology Faculdade de Ciências Médicas da Santa Casa de São Paulo (FCMSCSP), São Paulo, Brazil

**Keywords:** Obesity, Weight loss, Sibutramine, Topiramate, Anti-Obesity Agents

## Abstract

**Introduction:** Obesity is a chronic, prevalent, multifactorial disease, stigmatized and linked to multiple long-term complications, which makes the treatment currently available below of the expectations from what is necessary to combat it. Because of this limitation, off-label medications for the treatment of obesity, such as topiramate, have been widely used, in association with on-label medications, such as sibutramine, in order to achieve more effective weight loss.

**Objective:** Compare the effectiveness of the combination of sibutramine and topiramate with sibutramine monotherapy in weight loss in obese adults in regular follow-up. An effective weight loss is expected in the association group, due to complementary mechanisms of act, and antagonistic side effects of drugs, leading to greater weight loss and therapeutic adherence.

**Materials and Methods:** Retrospective cohort study with analysis of 66 medical records, distributed equally between groups. A weight loss of ≥ 5% was considered as an effectiveness criterion.

**Results:** The group treated with the association, after a mean follow-up period of 52 weeks, had a mean weight loss of 9.0%±8.2, against 5.3% ± 4.7 for the monotherapy group (p= 0.011). In the analysis of subgroups by weight loss categories, patients treated with the combination had an odds ratio of 6,7, to achieve ≥15% weight loss than the monotherapy group (OR=6.7, 95%CI=[1, 3; 33.7]).

**Conclusion:** A more effective weight loss was observed in the group treated with the combination of sibutramine and topiramate, when compared to the monotherapy group, after 52 weeks.

## INTRODUCTION

Obesity is a chronic disease associated with the excess of body fat due to an imbalance in energy balance (1–4) . It is known that excess adipose tissue, especially ectopic, is linked to increased production of pro-inflammatory cytokines and adipokines responsible for the development of comorbidities associated with weight gain, such as diabetes mellitus, hypertension, dyslipidemia, and some types of cancers (5–7). The treatment of obesity involves not only purely aesthetic goals but also the prevention, treatment, and control of comorbidities associated with it.

To achieve weight loss, maintaining a caloric deficit is crucial. However, obesity management involves more than the simple equation of consuming fewer calories and expending more energy, as the etiopathogenesis of weight gain is complex, encompassing genetic, sociocultural, behavioral, and biological causes. Biological aspects, such as the imbalance in homeostatic eating and reward system, along with peripheral and central signaling, constitute some of the elements explaining weight gain (5,8). As a complicating factor, the “set point” theory suggests a physiological inclination to return to the initial weight by reducing hormones related to satiety and energy expenditure, and increasing the desire for calorically dense foods (2,9–11). These mechanisms contribute to the high rate of weight regain, making treatment challenging and often frustrating for patients.

Against many factors that induce weight gain, pharmacological treatment becomes indispensable in the majority of cases. Sibutramine, a drug synthesized in the 1980s primarily for antidepressant purposes, was approved as an anti-obesity drug in 1997 due to its effect on satiety. It is a selective inhibitor of the reuptake of monoamines such as norepinephrine, serotonin, leading to increased levels of these neurotransmitters in appetite-regulating center, acting to reduce caloric intake (12–14) .

Associated with the increase in norepinephrine, some sympathomimetic side effects are expected, with the most common being insomnia, irritability, constipation, headache, and dry mouth (12,13). In 2010, the Sibutramine Cardiovascular Outcomes Trial (SCOUT), aimed to evaluate the impact of weight loss with sibutramine on the incidence of new cardiovascular events and death in individuals at very high risk. The primary outcome showed a 16% increase in the risk of non-fatal myocardial infarction in the sibutramine group compared to the placebo, mainly in those with high or very high cardiovascular risk. However, the same result was not found in the group without these risk factors (15). Such a study led to the prohibition of the medication in some countries in Europe (16). In Brazil, its use is authorized in patients under 65 years old, without established or suspected cardiovascular disease, and with controlled blood pressure levels (17).

Another medication used in clinical practice for obesity treatment is topiramate. For a long time, topiramate has been approved for use in epilepsy and migraine treatment due to mechanisms of action such as sodium channel blockade, kainate/AMPA receptor inhibition, carbonic anhydrase isoenzyme inhibition, and increased neurotransmitter gamma-aminobutyric acid (GABA) and glutamate (18). However, since 2002, after reports of weight loss in patients treated with topiramate, it has been used off-label as an anti-obesity drug.

The exact mechanism of action by which topiramate induces weight loss is not fully elucidated. It is believed that topiramate is associated with central and peripheral actions in reducing body weight. Among the possible central actions, topiramate would act on the arcuate nucleus, modulating GABA and glutamate receptors in the lateral hypothalamus, inhibiting orexigenic pathways (19–21). In the mesolimbic pathway, topiramate would modulate the brain reward circuit and synaptic plasticity by inhibiting the ventral tegmental area (VTA) through the release of GABAergic neurotransmitters and antagonizing AMPA/kainate glutamate receptors. This leads to a reduction in dopamine release in the nucleus accumbens, inhibiting addiction and compulsive behavior (22,23).

Among the side effects associated with the use of this anticonvulsant, the main ones are distal paresthesias and neuropsychiatric alterations such as cognitive impairment and slowing of thought processes, drowsiness, difficulty finding words, limiting its use as monotherapy. Additionally, topiramate is associated with midline defects during embryogenesis, being category D in pregnancy (24).

This retrospective study aims to compare the therapeutic efficacy between the isolated use of sibutramine and the combination of sibutramine with topiramate, with the expectation that combined use leads to greater weight loss than sibutramine monotherapy. This analysis was conducted based on a database of patients treated at a teaching hospital in São Paulo.

We believe that the combination is linked to acting on different points in the pathophysiology of obesity. While sibutramine concentrates its action on the homeostatic control of food intake, providing satiety, topiramate, on the other hand, acts on hedonic eating and the reward center. This dual approach can potentiate therapeutic benefits, encompassing different aspects of the complexity of obesity. Moreover, both present antagonistic side effects, which may assist in greater therapeutic adherence, reducing the individual side effects of each medication.

## METHODOLOGY

### Study Design

This is a retrospective study involving the analysis of medical record data from adult patients with obesity or overweight, treated at the Obesity Outpatient Clinic of the Endocrinology Department at Santa Casa de São Paulo from August 2016 to January 2022. Only patients who maintained regular follow-up for a minimum period of 52 weeks, corresponding to 3 medical appointments, were included, while those with discontinuous use of medications were excluded.

### Eligibility Criteria

Patients aged between 18 and 65 years with diagnoses of obesity and overweight and an indication for pharmacological treatment, according to the recommendation of the Brazilian Association for the Study of Obesity and Overweight (ABESO), were included. Obesity was considered for patients with a body mass index (BMI) ≥ 30 kg/m², and overweight was considered for those with BMI ≥ 27 kg/m². Criteria for pharmacological therapy indication were obesity or overweight with risk factors, after failure of non-pharmacological treatment (25).

Patients were excluded if they had:

- A history of type 2 diabetes with at least one other cardiovascular risk factor such as hypertension, dyslipidemia, current smoking, diabetic nephropathy with evidence of microalbuminuria.
- A history of coronary artery disease (angina or myocardial infarction), congestive heart failure, tachycardia, peripheral obstructive arterial disease, arrhythmia, or cardiovascular disease (stroke or transient ischemic attack), inadequately controlled hypertension (>145/90 mmHg).
- Use of other medications for obesity treatment or treatment of psychiatric disorders such as fluoxetine, sertraline.
- Personal or family history of nephrolithiasis.
- Personal history of closed-angle glaucoma.
- Pregnant or lactating individuals.

### Primary and Secondary Outcomes

Data including age, gender, history of anti-obesity medication treatment, sedentary lifestyle, and comorbidities associated with metabolic syndrome were collected.

Throughout the 3 consultations, the following information was collected:

- Weight (kg), weight loss (%), BMI (kg/m²)
- Systolic blood pressure (mmHg) and diastolic blood pressure (mmHg)
- Lipid profile: triglycerides (mg/dL), HDL-cholesterol (mg/dL), LDL-cholesterol (mg/dL)
- Glycemic profile: glycated hemoglobin (%)
- Side effects reported during the consultation
- Follow-up time (months)
- Medications dose (mg)

The primary outcome was evaluated based on the achieved weight loss, comparing the weight at the last medical evaluation to the weight at the beginning of the follow-up, for both applied therapies. A weight loss of ≥5% was considered effective, according to the FDA efficacy criteria for anti-obesity medications (26). Subsequently, patients were subdivided into 4 subgroups according to the percentage of weight loss: (1) <5%, (2) between 5 and 10%, (3) between 10 and 15%, (4) greater than 15%.

As a secondary outcome, the study assessed (1) the reduction in reported side effects in the sibutramine/topiramate group compared to monotherapy, and (2) the comparison of blood pressure, glycemic indices, lipid profiles, and renal parameters between the samples at the first and last consultations.

### Dosages

Doses of 10 or 15 mg of sibutramine and 25, 50, 75, and 100 mg of topiramate were used, titrated based on reported side effects during consultations. If there were no side effects, the doses were gradually increased to a maximum of 15 mg for sibutramine and/or 100 mg for topiramate. In case of mild to moderate side effects, the patient had their dose reduced to the lower dosage used previously, with the possibility of a new increase being reassessed in the following consultation. In the case of side effects considered severe, the medication was discontinued, and another therapy was introduced, thus being excluded of the study.

### Statistical Analysis

Statistical analysis was performed using Minitab 18.1, considering a level of statistical significance (α) of 5% for a two-tailed test.

In general, the t-student test was applied for independent quantitative samples, and the chi-square test for categorical parameters. Specifically, for the comparison of topiramate dosage between initial, intermediate, and final evaluations, the non-parametric Friedman test was used.

For the comparison of weight, weight loss, BMI, blood pressure, and laboratory parameters between groups, over the 3 evaluations, the analysis of variance (ANOVA) model with two factors (“group” and “evaluation”) was used, and whenever necessary, the Tukey multiple comparisons method was applied.

To calculate the odds ratio for weight loss in the sibutramine with topiramate group compared to the sibutramine group, logistic regression was used, with the weight loss parameter dichotomized for patients with loss ≥ 15% versus loss < 15%.

### Ethical Evaluation

All research subjects were studied according to the principles of the Declaration of Helsinki, the Nuremberg Code, and the Standards for Research Involving Human Beings (CNS Resolution 196/96) of the National Health Council, after project approval by the Research Ethics Committee of the Irmandade da Santa Casa de Misericórdia de São Paulo.

## RESULTS

A total of 118 patients who initiated the use of sibutramine (S) and/or combination (ST) between August 2016 and January 2021 were selected. Out of these patients, 25 were excluded from data collection due to therapy change, inability to afford medications, and intolerance to treatment after the first evaluation, leaving 45 patients using sibutramine with topiramate and 48 using sibutramine alone. Of these participants, 27 were excluded (12 in the combination group and 15 in the monotherapy group) due to a follow-up time shorter than proposed in the study, loss to follow-up, and poor adherence to treatment, resulting in 33 patients with regular follow-up for each group. (FIGURE 1)

**Figure 1.**
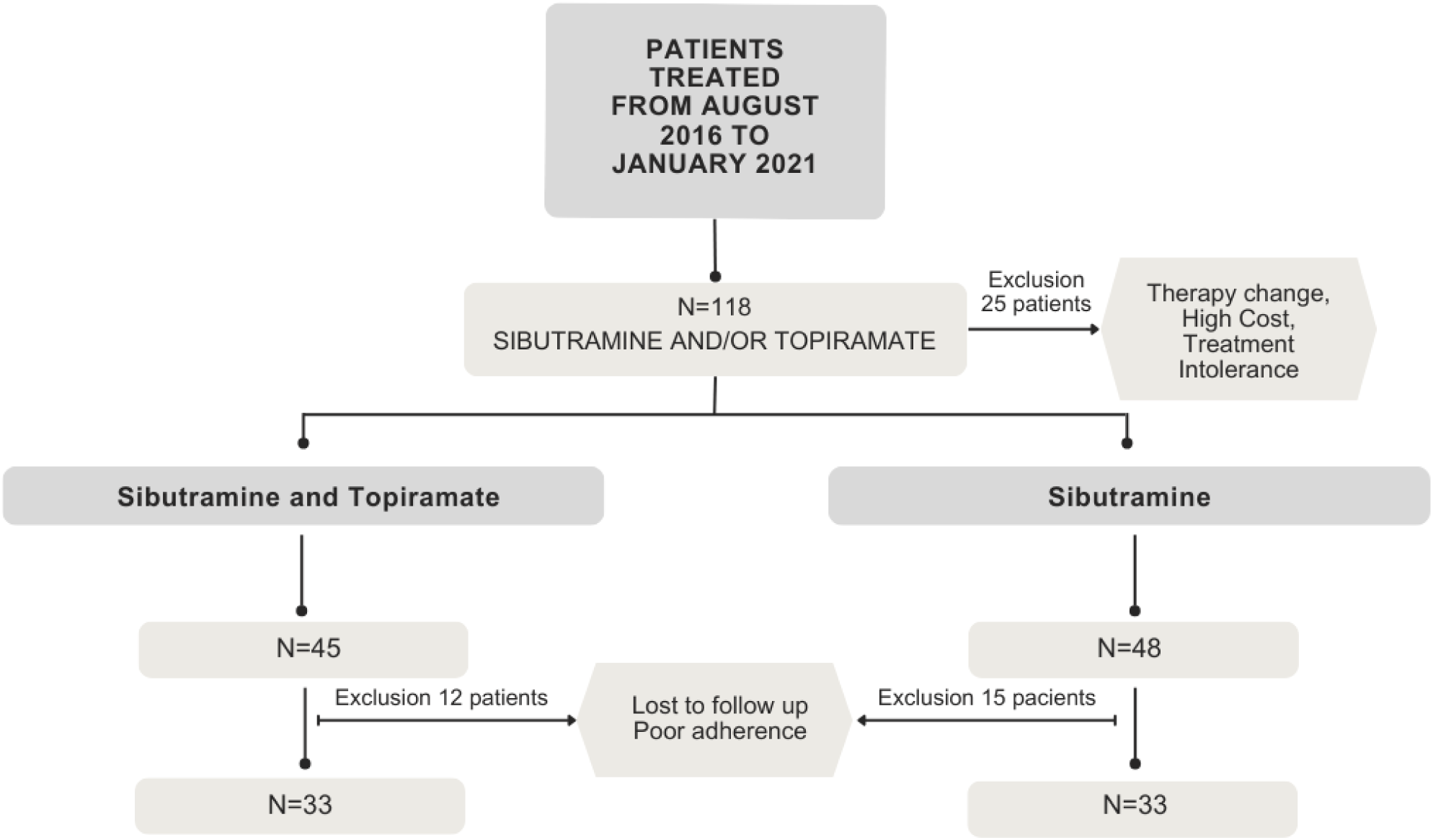
Sampling method

### Initial General Characteristics

The mean age was 42.6 years, ranging from 19 to 65 years, with the vast majority being female (78.8%). Regarding comorbidities at the beginning of treatment, 28.8% had a diagnosis of hypertension, 56.1% had prediabetes or type 2 diabetes, and 68.2% had dyslipidemia, with homogeneity between the groups. Despite the complications associated with obesity, less than half (43.1%) engaged in at least 150 minutes of physical activity per week. Moreover, when questioned about the history of medication treatment for obesity, only 30.3% in the sibutramine group had undergone any type of therapy, compared to 66.7% in the sibutramine with topiramate group, indicating a greater severity or refractoriness in the latter group. (Table 1)

**Table 1.** Characteristics of patients before treatment.

| **Table 1** | **Characteristics of patients before treatment** |  |  |  |
| --- | --- | --- | --- | --- |
|  | **Sibutramine** | **Sibutramine/Topiramate** | **Total** | **p-value** |
| **Age (y)** | 44,6±13 | 40,6±12,3 | 42,6±12,7 | 0,522<sup>1</sup> |
| **Female (%)** | 26 (78,8) | 26 (78,8) | 52 (78,8) |  |
| **Prior Treatment (%)** | 10 (30,3) | 22 (66,7) | 32 (48,5) | 0,003<sup>2</sup> |
| **Hypertension (%)** | 9 (27,3) | 10 (30,3) | 19 (28,8) | 0,786<sup>2</sup> |
| **IFG/Diabetes (%)** | 20 (60,6) | 17 (51,5) | 37 (56,1) | 0,457<sup>2</sup> |
| **Dyslipidemia (%)** | 24 (72,7) | 21 (63,6) | 45 (68,2) | 0,428<sup>2</sup> |
| **Sedentary Behavior** | 19 (59,4) | 18 (54,6) | 37 (56,9) | 0,694<sup>2</sup> |
| **Initial Weight (kg)** |  |  |  |  |
| Mean | 103,2±27,7 | 103,7±19,8 | 103,5±23,9 | 0,934<sup>1</sup> |
| Min-Max | 66,5-183 | 74,6-155 | 66,5-183 |  |
| **BMI (kg/m<sup>2</sup>)** |  |  |  |  |
| Mean | 39±6,2 | 39,3±5,4 | 39,1±5,8 | 0,828<sup>1</sup> |
| Overweight (%) | 2 (6,1) | 1 (3,0) | 3 (4,6) | 0,294<sup>2</sup> |
| Obesity Grade I (%) | 5 (15,2) | 7 (21,2) | 12 (18,2) |  |
| Obesity Grade II (%) | 16 (48,5) | 10 (30,3) | 26 (39,4) |  |
| Obesity Grade III (%) | 10 (30,3) | 15 (45,5) | 25 (37,9) |  |

BMI-Body Mass Index; IFG-Impaired fasting glucose; Min -mininum; Max-maximum; Y-years;
<sup>1</sup>t-student test; <sup>2</sup>chi-square test

### Initial Anthropometric Data

Regarding anthropometric data, the groups were quite similar. The overall mean weight was 103.5 ± 23.9 kg, and the BMI was 39.1 ± 5.8 kg/m2, indicating that the majority of patients had obesity grade II or III. In total, there were 3 cases (4.6%) with overweight, 12 (18.2%) with obesity grade I, 26 (39.4%) with obesity grade II, and 25 (37.9%) with obesity grade III. (Table 1)

### Initial Clinical and Laboratory Data

Regarding initial clinical data, a statistically significant difference was observed only in blood pressure, which was slightly higher in the Sibutramine with Topiramate group than in the Sibutramine alone group. As for laboratory parameters such as creatinine, total cholesterol and fractions, and glycated hemoglobin, there was no statistical difference between the groups. (Table 2)

**Table 2.**
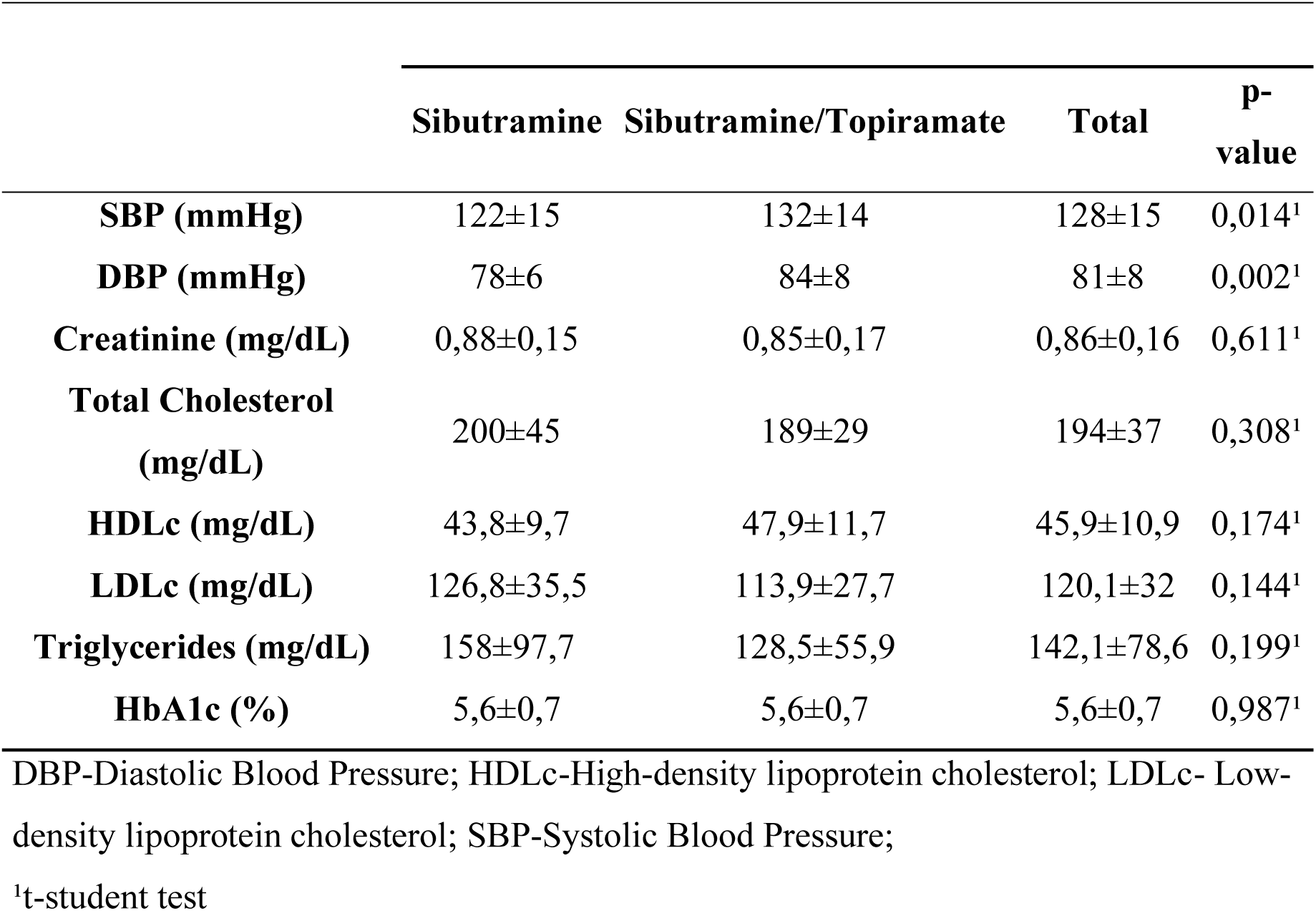
Initial clinical and laboratory data.

| **Table 2** | **Initial clinical and laboratory data** |  |  |  |
| --- | --- | --- | --- | --- |
|  | **Sibutramine** | **Sibutramine/Topiramate** | **Total** | **p-value** |
| **SBP (mmHg)** | 122±15 | 132±14 | 128±15 | 0,014<sup>1</sup> |
| **DBP (mmHg)** | 78±6 | 84±8 | 81±8 | 0,002<sup>1</sup> |
| **Creatinine (mg/dL)** | 0,88±0,15 | 0,85±0,17 | 0,86±0,16 | 0,611<sup>1</sup> |
| **Total Cholesterol (mg/dL)** | 200±45 | 189±29 | 194±37 | 0,308<sup>1</sup> |
| **HDLc (mg/dL)** | 43,8±9,7 | 47,9±11,7 | 45,9±10,9 | 0,174<sup>1</sup> |
| **LDLc (mg/dL)** | 126,8±35,5 | 113,9±27,7 | 120,1±32 | 0,144<sup>1</sup> |
| **Triglycerides (mg/dL)** | 158±97,7 | 128,5±55,9 | 142,1±78,6 | 0,199<sup>1</sup> |
| **HbA1c (%)** | 5,6±0,7 | 5,6±0,7 | 5,6±0,7 | 0,987<sup>1</sup> |

DBP-Diastolic Blood Pressure; HDLc-High-density lipoprotein cholesterol; LDLc- Low-density lipoprotein cholesterol; SBP-Systolic Blood Pressure;
<sup>1</sup>t-student test

### Treatment Dosages

At the beginning of treatment, approximately 78.8% were using the 10mg dose of sibutramine, progressing to the 15mg dose by the end of the total assessment, with 69.7% of patients using the maximum dose of 15mg. However, in the combination group, about 75.8% started treatment with 15mg of sibutramine, reaching 84.9% in the combination group by the end of the study. Despite this, there was no significant difference between the samples in the use of the 10mg or 15mg dose of sibutramine in the final assessment (p=0.142).

Regarding topiramate, it is observed, in general, that the administration of doses of 25mg and 50mg decreased, while doses of 75mg and 100mg increased from the initial to the final evaluation. In the final evaluation, 3% were using 25mg of topiramate, 36.4% were using 50mg, 15.2% were using 75mg, and 45.5% were using 100mg.

Considering the final percentage weight loss compared to the initial weight, a statistically significant difference was found between the groups (p=0.011). In the sibutramine and topiramate group, there was an average reduction of 6.7% and 9.0% in weight in the intermediate and final evaluations, while in the sibutramine group, it was 3.5% and 5.3% in the same evaluations. It was observed that some patients experienced weight gain, with 4 in the sibutramine group and 2 in the sibutramine with topiramate group at the intermediate evaluation, and another 6 at the final evaluation (3 in each group). (GRAPHIC 1)

**Graphic 1.**
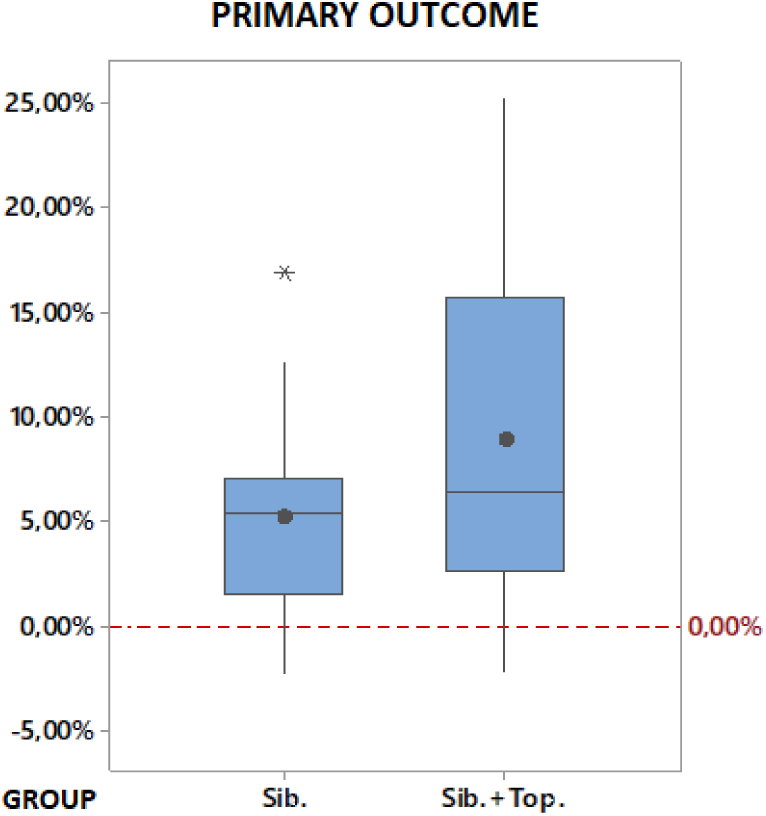
Primary outcome: ANOVA comparison between groups: p=0,011; interaction effect: p=0,647; Sib (Sibutramine); Sib+Top (Sibutramine plus Topiramate)

When assessing the weight loss categories, there was an almost 7-fold reduction in the chance of the sibutramine with topiramate group achieving a weight loss ≥ 15% compared to the sibutramine group (OR=6.7, 95% CI = [1.3; 33.7]), with 2 participants (6.1%) in the Sibutramine group versus 10 (30.3%) in the Sibutramine + Topiramate group. Due to the low casuistry in some cases, the statistical comparison between the groups was made with the following weight loss classes: “≤ 4.9%”, “5 to 9.9%”, and “≥ 10%”. At the intermediate evaluation, a statistically significant difference was observed between the groups with greater weight loss in the Sibutramine + Topiramate group (p=0.007), and at the final evaluation, the Sibutramine + Topiramate group also showed greater weight loss, although not statistically significant (but very close to statistical significance), with p=0.056.

Regarding reported side effects at the beginning of treatment, a total of 5 patients (15.15%) in the sibutramine group reported side effects such as restlessness, insomnia, constipation, nausea, and palpitation, compared to 10 (30.30%) in the combination group reporting restlessness, irritability, insomnia, paresthesias, and drowsiness. At the end of the study, there was a reduction in the number of complaints to 2 in the sibutramine group and 5 in the topiramate group. However, this difference was not statistically significant (p=0.159). No serious adverse events were reported during the follow-up.

There was no statistically significant difference between the groups for any of the clinical and laboratory parameters evaluated. Regarding systolic and diastolic blood pressure, they were the only parameters that showed a different evolution (interaction effect). It was observed that in the Sibutramine group, there is a tendency for an increase, while in the combination group, there is a tendency for a decrease in pressure between the initial and final evaluation (significant interaction, with p=0.033 for systolic blood pressure, and p=0.041 for diastolic blood pressure), but still, without a significant difference between the groups and visits. For both groups, there was a significant reduction in total cholesterol and LDL, but there was no difference between the groups.

## DISCUSSION

The retrospective study demonstrated that the combination of sibutramine with topiramate is associated with greater effectiveness in weight loss compared to monotherapy with sibutramine, corresponding to a weight loss of 9.0% versus 5.3%, respectively, with an absolute mean weight loss of -9.5kg and -5.2kg. A literature review was conducted on major platforms such as PubMed and Scielo, however, no other similar studies with the same comparison between medications were found. When analyzed individually, in a similar treatment period, isolated sibutramine at a dose of 15mg led to a weight loss of -4.2kg (95% CI=[-5.73 to -2.28]), similar to what was found in our study (27–30). Regarding isolated topiramate, a meta-analysis assessed the efficacy and safety of the medication in clinical trials lasting more than 28 weeks, identifying an average weight loss of 6.58kg (95% CI=[-6.12 to -4.56kg]) (31–35). Therefore, the combination showed greater weight loss compared to the isolated use of the medications, even in other studies in the literature.

When analyzing the previous history of drug treatment for obesity, a higher number of patients was observed in the combination group, as well as a greater tendency to introduce the dose of 15mg of sibutramine, already in the first intervention, due to previous attempts at drug treatment, and in some cases, even with sibutramine. Nevertheless, there was no difference between the doses of sibutramine applied until the last assessment. It was observed that in the sibutramine with topiramate group, there was a more accelerated weight loss until the second intervention, with a deceleration of weight loss afterward. In the sibutramine group, on the other hand, weight loss continued steadily until the last assessment. However, when stratifying each group by the percentage of weight loss achieved at the end of the assessment, a odds ratio of 6.7 (OR=6.7, 95% CI=[1.3; 33.7]) in achieving a weight loss greater or equal to 15% than in monotherapy was evident.

Among the possible justifications for greater weight loss with dual therapy is the action at different points in the physiology of obesity, operating complementarily. Sibutramine, acting on the inhibition of serotonin and noradrenaline reuptake, leads to increased satiety and reduced caloric intake (36–38). Topiramate, however, does not have a fully elucidated mechanism of action for weight loss. Acting on different biochemical pathways, such as voltage-dependent sodium and GABA receptors, increasing glutamate through kainate/NMDA receptors, and inhibiting carbonic anhydrase, much is postulated about the action of this medication (18,24).

Various studies have linked the action of topiramate in weight loss, not only through the reduction of appetite by acting on caloric intake but also through the inhibition of the brain reward center. It is known that glutamatergic and GABAergic receptors are located at different points in the central nervous system, such as the hypothalamus, both being linked to the regulation of food intake (19). Studies with patients with binge eating disorder, where there is an imbalance in the mesolimbic dopaminergic system, have obtained promising results with topiramate due to its action as a GABAergic agonist and glutamatergic antagonist, inhibiting the brain reward system (23,39). It is known that about 32.8–41.7% of obese patients present binge eating, perhaps justifying the greatest weight loss in the association group (40). Thus, we believe that the synergistic action of sibutramine and topiramate at different points in the pathophysiology of obesity influenced the greater weight loss in the combination group.

Regarding secondary outcomes, literature data differ regarding the benefit of topiramate on blood glucose and lipids. Some studies have correlated topiramate with an increase in adiponectin, consequently leading to improved insulin sensitivity, lipid and free fatty acid oxidation. Other studies speculate a correlation between topiramate and increased insulin secretion. However, more studies are needed, questioning whether this effect would not be a direct consequence of weight loss by the medication (35,41,42). In the present study, no difference was found in glycemic and lipid parameters, although both groups showed a reduction in total cholesterol and LDL similarly.

Throughout the study, there was a tendency for a decrease in systolic and diastolic blood pressure in the combination group compared to monotherapy, where there was a tendency for an increase. In the SCOUT study, there was a 16% increased cardiovascular risk for patients using sibutramine compared to placebo, linked to an increase in blood pressure and heart rate (15). Topiramate, on the other hand, has been related to a reduction in blood pressure, albeit through unknown mechanisms (34). However, in the present study, there was no statistical significance regarding blood pressure reduction. Since it is a non-interventional study, it was not possible to exclude possible confounding biases, as many patients started treatment with antihypertensives, statins, and hypoglycemics throughout follow-up. In addition, despite most of the initial characteristics among participants being equal regarding blood pressure, there was no homogeneity between the samples, being higher in the combination group.

Regarding side effects, the main ones reported were agitation, tachycardia, xerostomia, constipation, nausea, and malaise in the group treated only with sibutramine. For the combination group, there were also paresthesias, drowsiness, forgetfulness, and dizziness. In the final assessment, there was a reduction in reported complaints. Due to the low casuistry, it was not possible to identify a significant difference between the groups. Information bias associated with heterogeneity between medical records was an important interfering factor in collecting this data.

The retrospective study indicates that the combination of sibutramine and topiramate exhibits superior efficacy in weight reduction compared to sibutramine monotherapy. Individual analysis of the medications yielded results consistent with existing literature, highlighting the positive impact of both on weight loss. The absence of comparable studies in the literature emphasizes not only the uniqueness of our analysis but also the necessity for more robust studies, such as controlled clinical trials, to validate and elucidate these findings.

## CONCLUSION

Similar weight loss was observed in the group treated with the combination of sibutramine and topiramate compared to the group treated only with sibutramine after a mean follow-up period of 52 weeks. It was not possible to assess secondary outcomes, such as improvements in metabolic or cardiovascular parameters.

## Data Availability

All data produced in the present study are available upon reasonable request to the authors

## ACKNOWLEDGMENTS

Disclosure: no potential conflict of interest relevant to this article was reported.

## REFERENCES

1. Mechanick JI, Garber AJ, Handelsman Y, Garvey WT, Beir DM, Bohannon NJV, et al. American Association of Clinical Endocrinologists’ Position Statement on Obesity and Obesity Medicine. Endocr Pract. 2012 Sep;18(5):642–8. doi:10.4158/EP14280.PS

2. Garvey WT. Is Obesity or Adiposity-Based Chronic Disease Curable: The Set Point Theory, the Environment, and Second-Generation Medications. Endocr Pract. 2022 Feb;28(2):214–22. doi 10.1016/j.eprac.2021.11.082.

3. Jastreboff AM, Kotz CM, Kahan S, Kelly AS, Heymsfield SB. Obesity as a Disease: The Obesity Society 2018 Position Statement. Obesity. 2019 Jan;27(1):7–9. doi:10.1002/oby.22378.

4. Yumuk V, Tsigos C, Fried M, Schindler K, Busetto L, Micic D, et al. European Guidelines for Obesity Management in Adults. Obes Facts. 2015;8(6):402–24. doi: 10.1159/000442721.

5. Heymsfield SB, Wadden TA. Mechanisms, Pathophysiology, and Management of Obesity. N Engl J Med. 2017 Jan 19;376(3):254–66. doi: 10.1056/NEJMra1514009.

6. Ryan DH, Yockey SR. Weight Loss and Improvement in Comorbidity: Differences at 5%, 10%, 15%, and Over. Curr Obes Rep. 2017 Jun;6(2):187–94. doi: 10.1007/s13679-017-0262-y.

7. Bray GA, Heisel WE, Afshin A, Jensen MD, Dietz WH, Long M, et al. The Science of Obesity Management: An Endocrine Society Scientific Statement. Endocr Rev. 2018 Apr 1;39(2):79–132. doi: 10.1210/er.2017-00253.

8. Oussaada SM, Van Galen KA, Cooiman MI, Kleinendorst L, Hazebroek EJ, Van Haelst MM, et al. The pathogenesis of obesity. Metabolism. 2019 Mar;92:26–36. doi: 10.1016/j.metabol.2018.12.012.

9. Ochner CN, Barrios DM, Lee CD, Pi-Sunyer FX. Biological mechanisms that promote weight regain following weight loss in obese humans. Physiol Behav. 2013 Aug;120:106–13. doi: 10.1016/j.physbeh.2013.07.009.

10. Van Baak MA, Mariman ECM. Mechanisms of weight regain after weight loss — the role of adipose tissue. Nat Rev Endocrinol. 2019 May;15(5):274–87. doi: 10.1038/s41574-018-0148-4.

11. MacLean PS, Higgins JA, Giles ED, Sherk VD, Jackman MR. The role for adipose tissue in weight regain after weight loss. Obes Rev. 2015 Feb;16(S1):45–54. doi: 10.1111/obr.12255.

12. McNeely W, Goa KL. Sibutramine: A Review of its Contribution to the Management of Obesity. Drugs. 1998;56(6):1093–124. doi: 10.2165/00003495-199856060-00019.

13. Nisoli E, Carruba MO. An assessment of the safety and efficacy of sibutramine, an anti-obesity drug with a novel mechanism of action. Obes Rev. 2000 Oct;1(2):127–39. doi: 10.1046/j.1467-789x.2000.00020.x.

14. Lean M. How does sibutramine work? Int J Obes. 2001 Dec;25(S4):S8–11. doi: 10.1038/sj.ijo.0801931.

15. Caterson ID, Finer N, Coutinho W, Van Gaal LF, Maggioni AP, Torp-Pedersen C, et al. Maintained intentional weight loss reduces cardiovascular outcomes: results from the Sibutramine Cardiovascular OUTcomes (SCOUT) trial. Diabetes Obes Metab. 2012 Jun;14(6):523–30. doi: 10.1111/j.1463-1326.2011.01554.x.

16. 16. European Medicines Agency (EMA). European Medicines Agency recommends suspension of marketing authorisation for sibutramine [Internet]. 2010 [cited 2024 Jan 20]. Available from: https://www.ema.europa.eu/en/news/european-medicines-agency-recommends-suspension-marketing-authorisation-sibutramine?fbclid=IwAR0j-JZEdrJ3qbW2iDOGiVJoiIPZVKuJSZkV5j9a438Lt9h6O6lMP0c34qA

17. Agencia Nacional de Vigilancia Sanitária (ANVISA). Resolução - RDC n° 52, de 6 de outubro de 2011. RDC. Sect. 1, 52 Oct 6, 2011.

18. De Simone G, Supuran C. Antiobesity Carbonic Anhydrase Inhibitors. Curr Top Med Chem. 2007 May 1;7(9):879–84. doi: 10.2174/156802607780636762

19. Da Silva AA, Campanella LCA, Ramos MC, Faria MS, Paschoalini MA, Marino-Neto J. Ingestive effects of NMDA and AMPA-kainate receptor antagonists microinjections into the lateral hypothalamus of the pigeon (Columba livia). Brain Res. 2006 Oct;1115(1):75–82. doi: 10.1016/j.brainres.2006.07.073.

20. Stanley BG, Urstadt KR, Charles JR, Kee T. Glutamate and GABA in lateral hypothalamic mechanisms controlling food intake. Physiol Behav. 2011 Jul;104(1):40–6. doi: 10.1016/j.physbeh.2011.04.046.

21. Rossi MA, Basiri ML, McHenry JA, Kosyk O, Otis JM, Van Den Munkhof HE, et al. Obesity remodels activity and transcriptional state of a lateral hypothalamic brake on feeding. Science. 2019 Jun 28;364(6447):1271–4. doi: 10.1126/science.aax1184.

22. Turner BD, Kashima DT, Manz KM, Grueter CA, Grueter BA. Synaptic Plasticity in the Nucleus Accumbens: Lessons Learned from Experience. ACS Chem Neurosci. 2018 Sep 19;9(9):2114–26. doi: 10.1021/acschemneuro.7b00420.

23. Himmerich H, Treasure J. Psychopharmacological advances in eating disorders. Expert Rev Clin Pharmacol. 2018 Jan 2;11(1):95–108. doi: 10.1080/17512433.2018.1383895.

24. McElroy SL, Guerdjikova AI, Martens B, Keck PE, Pope HG, Hudson JI. Role of Antiepileptic Drugs in the Management of Eating Disorders: CNS Drugs. 2009;23(2):139–56. doi: 10.2165/00023210-200923020-00004.

25. 25. Associação Brasileira para o Estudo da Obesidade e da Sindrome Metabolica (ABESO). Diretrizes brasileiras de obesidade 2016. São Paulo, SP: ABESO; 2016. Report No.: 4^a^ edição. [cited 2024 Jan 20]. Available from: http://abeso.org.br/diretrizes/

26. Center for Drug Evaluation and Research. Guidance for Industry Developing Products for Weight Management. 2007 Feb;

27. James WPT, Astrup A, Finer N, Hilsted J, Kopelman P, Rössner S, et al. Effect of sibutramine on weight maintenance after weight loss: a randomised trial. The Lancet. 2000 Dec;356(9248):2119–25. doi: 10.1016/s0140-6736(00)03491-7.

28. Apfelbaum M, Vague P, Ziegler O, Hanotin C, Thomas F, Leutenegger E. Long-term maintenance of weight loss after a very-low-calorie diet: a randomized blinded trial of the efficacy and tolerability of sibutramine. Am J Med. 1999 Feb;106(2):179–84. doi: 10.1016/s0002-9343(98)00411-2.

29. 29. Mathus-Vliegen EMH, for the Balance Study Group. Long-term maintenance of weight loss with sibutramine in a GP setting following a specialist guided very-low-calorie diet: a double-blind, placebo-controlled, parallel group study. Eur J Clin Nutr. 2005 Aug 1;59(S1):S31–9. doi: 10.1038/sj.ejcn.1602172.

30. Rucker D, Padwal R, Li SK, Curioni C, Lau DCW. Long term pharmacotherapy for obesity and overweight: updated meta-analysis. BMJ. 2007 Dec 8;335(7631):1194–9. doi: 10.1136/bmj.39385.413113.25.

31. Kramer CK, Leitão CB, Pinto LC, Canani LH, Azevedo MJ, Gross JL. Efficacy and safety of topiramate on weight loss: a meta-analysis of randomized controlled trials. Obes Rev. 2011 May;12(5):e338–47. doi: 10.1111/j.1467-789X.2010.00846.x.

32. Astrup A, Toubro S. Topiramate: A New Potential Pharmacological Treatment for Obesity. Obes Res. 2004 Dec;12(S12). doi: 10.1038/oby.2004.284.

33. Stenlöf K, Rössner S, Vercruysse F, Kumar A, Fitchet M, Sjöström L, et al. Topiramate in the treatment of obese subjects with drug-naive type 2 diabetes. Diabetes Obes Metab. 2007 May;9(3):360–8. doi: 10.1111/j.1463-1326.2006.00618.x.

34. Wilding J, Gaal LV, Rissanen A, Vercruysse F, Fitchet M. A randomized double-blind placebo-controlled study of the long-term efficacy and safety of topiramate in the treatment of obese subjects. Int J Obes. 2004 Nov;28(11):1399–410. doi: 10.1038/sj.ijo.0802783.

35. Eliasson B, Gudbjörnsdottir S, Cederholm J, Liang Y, Vercruysse F, Smith U. Weight loss and metabolic effects of topiramate in overweight and obese type 2 diabetic patients: randomized double-blind placebo-controlled trial. Int J Obes. 2007 Jul;31(7):1140–7. doi: 10.1038/sj.ijo.0803548.

36. Hansen DL, Toubro S, Stock MJ, Macdonald IA, Astrup A. Thermogenic effects of sibutramine in humans. Am J Clin Nutr. 1998 Dec;68(6):1180–6. doi: 10.1093/ajcn/68.6.1180.

37. Luque CA, Rey JA. Sibutramine: A Serotonin–Norepinephrine Reuptake-Inhibitor for the Treatment of Obesity. Ann Pharmacother. 1999 Sep;33(9):968–78. doi: 10.1345/aph.18319.

38. Heal DJ, Cheetham SC, Prow MR, Martin KF, Buckett WR. A comparison of the effects on central 5-HT function of sibutramine hydrochloride and other weight-modifying agents. doi: 10.1038/sj.bjp.0702067.

39. Reas DL, Grilo CM. Pharmacological treatment of binge eating disorder: update review and synthesis. Expert Opin Pharmacother. 2015 Jul 3;16(10):1463–78. doi: 10.1517/14656566.2015.1053465.

40. Kessler RC, Berglund PA, Chiu WT, Deitz AC, Hudson JI, Shahly V, et al. The Prevalence and Correlates of Binge Eating Disorder in the World Health Organization World Mental Health Surveys. Biol Psychiatry. 2013 May;73(9):904–14. doi: 10.1016/j.biopsych.2012.11.020.

41. Wilkes JJ, Nelson E, Osborne M, Demarest KT, Olefsky JM. Topiramate is an insulin-sensitizing compound in vivo with direct effects on adipocytes in female ZDF rats. Am J Physiol-Endocrinol Metab. 2005 Mar;288(3):E617–24. doi: 10.1152/ajpendo.00437.2004.

42. Wilkes JJ, Nguyen MTA, Bandyopadhyay GK, Nelson E, Olefsky JM. Topiramate treatment causes skeletal muscle insulin sensitization and increased Acrp30 secretion in high-fat-fed male Wistar rats. Am J Physiol-Endocrinol Metab. 2005 Dec;289(6):E1015–22. doi: 10.1152/ajpendo.00169.2005.

